# Identifying Dietary Consumption Patterns from Survey Data: A Bayesian Nonparametric Latent Class Model

**DOI:** 10.1101/2021.11.18.21266543

**Authors:** Briana J.K. Stephenson, Stephanie M. Wu, Francesca Dominici

## Abstract

**Summary:** Dietary intake is one of the largest contributing factors to cardiovascular health in the United States. Amongst low-income adults, the impact is even more devastating. Dietary assessments, such as 24-hour recalls, provide snapshots of dietary habits in a study population. Questions remain on how generalizable those snapshots are in nationally representative survey data, where certain subgroups are sampled disproportionately to comprehensively examine the population. Many of the models that derive dietary patterns account for study design by incorporating the sampling weights to the derived model parameter estimates post hoc. We propose a Bayesian overfitted latent class model that accounts for survey design and sampling variability. Compared to other standard approaches used for survey data, our model showed improved identifiablity of the true population prevalence and pattern distribution in simulation. Using dietary intake data from the 2011-2018 National Health and Nutrition Examination Survey, we demonstrated the utility of our model to derive dietary patterns in adults considered low-income (at or below the 130% poverty income threshold), to understand if and how these patterns generalize in a smaller sub-population. A total of five dietary patterns were identified and characterized. Reproducible code/data are provided on GitHub to encourage further research and application in this area.

## 1. Introduction

### 1.1. Motivation

The impact of poor diet has continually devastated the United States, accounting for over 500,000 deaths annually, with 84% of those deaths due to cardiovascular disease (CVD)(Mokdad et al., 2018; Roth et al., 2018). The negative health impacts of poor diet disproportionately affect low-income and racial minority populations (Brown et al., 2018; Fahlman et al., 2010). Understanding the dietary consumption behaviors that contribute to poor health in these target populations may help in tailoring interventions and resources to improve their nutritional health.

Through the implementation of complex survey designs and targeted recruitment strategies, researchers are able to obtain more representative population samples in an effort to better understand populations of greatest interest. Consequently, survey sampling methodologies have been developed to improve population-based estimates and generate appropriate inference based on the sampled data.

While low-income and racial minority adults are a population at greatest risk of poor diet and subsequently poorer health outcomes, they are often underrepresented in survey studies (Tourangeau et al., 2014). In an effort to achieve a more nationally representative sample, surveys such as the National Health and Nutrition Examination Survey (NHANES) have corrected for this underrepresentation by oversampling demographics of greater public health interest to improve the accuracy and reliability of national-based estimates of health outcomes and exposures (Zipf et al., 2013). Unfortunately, most of the current statistical approaches for deriving dietary patterns from survey data do not incorporate the weights during estimation, which could lead to biased and inconsistent data-driven patterns for population demographics.

Latent class models (LCM) are an effective tool to comprehensively analyze consumption patterns of a full set of foods included on a diet assessment (Sotres-Alvarez et al., 2010; Keshteli et al., 2015). Implementation of this procedure is available on commonly used statistical software such as SAS (Proc LCA) and R (poLCA) and offer parameter estimation of latent class model parameters via frequentist algorithms (e.g. Expectation-Maximization and Newton-Raphson) (Lanza et al., 2007; Linzer and Lewis, 2011; Muthén and Shedden, 1999). Bayesian estimation is accomplished through an R package (BayesLCA), but is limited to binary consumption responses (White and Murphy, 2014).

Patterns derived from LCA are dependent on the observed study data. This is of concern when the study data is not representative of the study population. For example, historically, certain subgroups of the population have been undersampled and underrepresented in studies. In other scenarios, surveys may purposely oversample subgroups to gain more information from them. Demographics that dominate in a population often mask dietary habits of smaller-sized demographics, which may deviate from the majority habit. Study designs have strived to correct for this through the implementation of sampling weights that account for underrepresnetation and nonresponse. However, none of these standard packages previously described incorporate sampling weights directly into the estimation procedures. Mplus is one of the few statistical softwares available to adjust for complex survey design, but is limited under a frequentist framework, which can present issues with matrix inversion and computational demand when handling the high-dimensionality of diet data, which can be large and sparse for rarely consumed food items (Muthé and Muthén, 2017).

### 1.2. Potential Solutions in Bayesian Nonparametrics

Bayesian nonparametrics offers a more efficient solution that is able to (1) accommodate the complex high dimensionality of dietary intake data, (2) handle large-sized populations, such as the United States, (3) reduce multiple model testing and fitting to determine the appropriate number of patterns, (4) preserve model stability in the presence of sparsely consumed foods, and (5) integrate prior information with observed data. These features improve parameter estimation and subsequent population inference (Hjort et al., 2010; Liu, 2008).

Bayesian survey data applications have centered mostly on generating inference for derived population-based estimates (Si et al., 2015; Savitsky and Toth, 2016; Gunawan et al., 2020), but have not been fully explored in regards to model-based clustering. Bayesian nonparametric mixture models that utilized dietary intake data either did not contain complex survey data (Fahey et al., 2007; De Vito et al., 2019; Stephenson et al., 2020a), or applied sampling weights posthoc after model parameter estimation was complete (Stephenson et al., 2020b; De Vito et al., 2022). Kunihama et al. (2016) is one of the few that introduced a sampling algorithm that incorporates survey weights directly into the estimation, but did not take into account sampling variability present in nationally-representative surveys.

Our overall objective is to examine the dietary patterns of low-income adults in the United States. This adult subpopulation represents a minority of the United States and the patterns of this demographic are often masked by the majority of adults not classified as low-income. We have built upon the survey sampling framework and added the following contributions: (1) implemented an overfitted latent class model, which is asymptotically similar to the Dirichlet Process mixture model; (2) extended and integrated the works of Kunihama et al. (2016) and Savitsky and Toth (2016) to generate population-based estimates that also adjust for sampling variability in the survey design; (3) demonstrated the utility of this approach by applying this model to publicly available national survey data to derive nationally representative dietary consumption patterns of low-income adults in the United States from 2011-2018; and (4) provided publicly available reproducible code for researchers to apply this technique on future national dietary survey data.

We organize this paper as follows: Section 2 describes our proposed weighted overfitted latent class model. Section 3 compares our model with current model-based approaches for survey data via a simulation study. Section 4 describes the National Health and Nutrition Examination Survey. Section 5 presents results of the method applied to the National Health and Nutrition Examination Survey. Section 6 discusses next steps and future directions.

## 2. Weighted Overfitted Latent Class Model

A weighted overfitted latent class model is a Bayesian nonparametric technique that can be used to identify subgroups or clusters within a survey sample that share common behaviors amongst a set of observed nominal variables (Van Havre et al., 2015). It can be seen as an extension of the latent class model, which typically requires multiple fits and post hoc testing to determine the appropriate number of latent classes or patterns. The overfitted latent class model removes this redundancy by overfitting the model with a large number of latent classes (or clusters) and allowing a data-driven approach to choosing the number of latent clusters. Empty clusters are able to drop out of the model during the Markov chain Monte Carlo Gibbs sampling algorithm, and nonempty clusters remain. Each participant is assigned to one of the derived clusters, corresponding to a dietary pattern. The overfitted structure is also asymptotically equivalent to the Dirichlet Process model, allowing additional flexibility within a Bayesian nonparametric framework (Van Havre et al., 2015).

### 2.1. Overfitted Latent Class Model (OLCM)

We define some notation of the standard latent class model, with a sampled population of size *n* and *K* unique dietary patterns, where each pattern describes the consumption of *p* food items. Let ***y***_***i·***_ = (*y*_*i*1_, …, *y*_*ip*_) denote the set of *p* observed food items. Each observed food item, *y*_*ij*_, is categorical, where *y*_*ij*_ ∈ {1, 2, …, *d*_*j*_} is individuals *i*’s consumption level for food item *j*. Let *π*_*k*_ denote the probability of assignment to dietary pattern *k* ∈ {1, …, *K*}, and *π* = (*π*_1_, …, *π*_*K*_). The dietary pattern assignment of individual *i* ∈ {1, …, *n*} from the sampled population is denoted by *z*_*i*_. Let *θ*_*jc*|*k*_ denote the probability of consuming food item *j*, at the *c* ∈ {1, …, *d*_*j*_} consumption level, given an individual’s assignment to diet pattern *k*, where *d*_*j*_ is the maximum consumption level for food item *j*, and *θ* = *{θ*_*jc*|*k*_}_*j,k*_. The subject-specific likelihood is then defined as

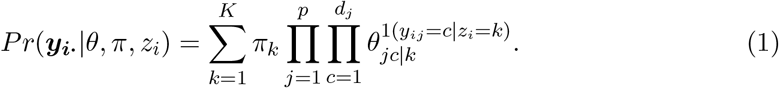

The likelihood for the overfitted latent class model shares the same structure as that of the standard latent class model shown in (1), but since *K* is typically not known in practice, it is fixed to an exceedingly large number that asymptotically simulates an infinite mixture model (Van Havre et al., 2015). Under a Bayesian estimation framework, the model parameters are updated in the Gibbs sampler based on the number of observed individuals classified to a given latent class or consumption level. For example, exploiting the convenience of conjugacy, the probability vector, ***π*** = (*π*_1_, …, *π*_*K*_), follows a Dirichlet prior and conditional posterior with hyperparameters for each latent class defined as (*α*_1_, …, *α*_*K*_):

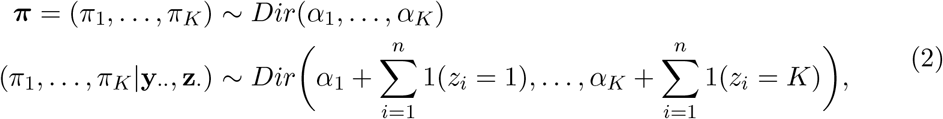

where **y**_*··*_ = (**y**_**1***·*_, …, **y**_**n***·*_) and ***z***_***·***_ = (*z*_1_, …, *z*_*n*_). With no prior knowledge on the number of classes, we utilize a noninformative, flat Dirichlet prior, where *α*_1_ = *α*_2_ = …= *α*_*K*_ = *α*. This hyperparameter moderates the rate of growth for nonempty latent classes. The smaller the hyperparameter, the slower nonempty clusters will form. Similarly, we assume no prior knowledge on the consumption pattern of each observed food, such that 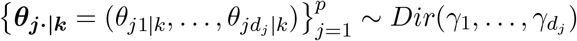 for all *k* ∈ {1, 2, …, *K*} is also fit with a non-informative flat Dirichlet prior with a constant *γ* hyperparameter (*γ*_1_ = *γ*_2_ = *γd_j_* = *γ*).

### 2.2. Extension to Weighted Overfitted Latent Class Model (wtOLCM)

Incorporating survey weights in a Bayesian setting serves as a natural extension to the overfitted latent class model. As described in Kunihama et al. (2016), information used to update each model parameter is enhanced with weights, simulating a pseudo-like population that is similar in size and structure to the target population. A normalization constant is used to ensure the weights sum to the target population. This enables dietary patterns to form in accordance with the target population, but does not consider changes that can occur in size and composition from one sampled population to another. Sampling variability should be considered in the model, and precision estimates should reflect the sample size rather than the population size. Otherwise, uncertainty surrounding model estimation will be biased. To address this limitation, we instead propose an approach similar to Savitsky and Toth (2016) and normalize the sampling weights to sum to the sample size. This will account for sampling variability while allowing model estimates to generalize better to the target population.

Let *w*_*i*_ denote the sampling weight of study participant *i* ∈ {1, …, *n*}. We impose fixed normalization constant, *κ*, where 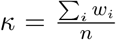, with *n* denoting the study sample size. With this newly defined *κ* and with ***w***_*·*_ = (*w*_1_, …, *w*_*n*_), the conditional posterior of the probability of assignment vector, ***π*** = (*π*_1_, …, *π*_*K*_), updates based on the weighted number of participants assigned to each pattern:

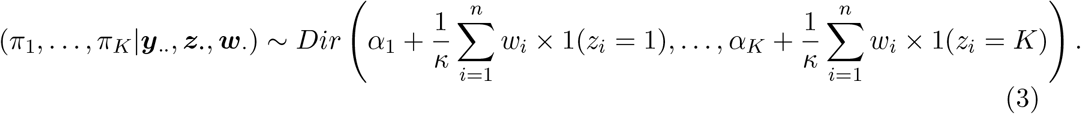

Similarly, for the consumption level distribution of each dietary pattern,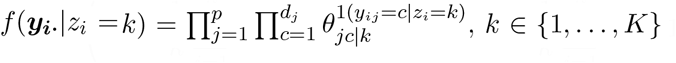, updates for the conditional posteriors of the probabilities of consumption are based on the weighted number of participants that share dietary consumption behaviors. For all *j* ∈ {1, …, *p*} and *k* ∈ {1, …, *K*},

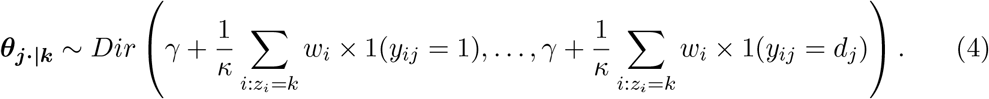

The full likelihood model is written as:

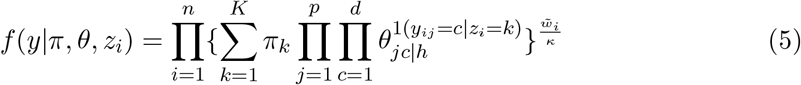

## 3. Simulation Study

### 3.1. Survey-weighted Approaches

We performed our simulation study under three different approaches for handling survey data. Method 1 serves as our control, a standard overfitted latent class model, where sample weights are ignored (unweighted OLCM). Method 2 provides an alternative approach, a weighted finite population Bayesian bootstrap (WFPBB) (Gunawan et al., 2020; Dong et al., 2014), where pseudo-representative samples are generated by using survey weights to “undo” the unequal sampling scheme and impute synthetic populations, and then approximate simple random samples are drawn from these synthetic populations. This application of the WFPBB method builds on earlier work of pseudo-population generation through multiple imputation techniques (Raghunathan et al., 2003; Zhou et al., 2016). Implementation details of this method are provided in Supplementary Section 1. Method 3 is our proposed weighted overfitted latent class model (wtOLCM) that extends the work of Kunihama et al. (2016) and Savitsky and Toth (2016) where the sample weights are directly incorporated into the sampling algorithm, as detailed in section 2. Our simulation study will evaluate how well these three methods are able to identify the true population prevalence of dietary patterns using the sampled data.

### 3.2. Simulation Setup

The goal of our simulation study is to compare how well the three methods are able to identify the true population prevalence, as well as the composition of the true population patterns. Algorithm run time is also compared for computational reference. We consider a simulated population of size *N* = 5000. A total of *K*_*true*_ = 3 patterns exist in the population with probability distribution ***π***_***true***_ = (0.1, 0.3, 0.6). Each pattern consists of *p* = 50 categorical variables that can take on values 1, 2, 3 or 4. For case A, the mode was set at 0.85 for the true pattern value of interest, and at 0.05 for the remaining three values. To evaluate under additional noise, case B was performed where the mode was set at 0.55, and 0.15 for all other remaining values. Pattern 1 was defined with a mode at level 3 for the first 25 variables, and a mode at level 1 for the remaining 25 variables. Pattern 2 was defined with a mode at level 2 for the first 10 variables, and a mode at level 4 for the remaining 40 variables. Pattern 3 was defined with a mode at level 1 for the first 10 variables, a mode at level 2 for the next 20 variables, and a mode at level 3 for the remaining 20 variables. Subjects were initially assigned to one of these three patterns, and the subject-specific observed data was simulated by drawing from a multinomial distribution for each of the 50 corresponding variables described above based on the assigned pattern. The total population was comprised of *S* = 4 disproportionate subpopulations containing varied distributions of the three patterns (Supplementary Table 1). A subset of 100 subjects were randomly selected from each of the simulated subpopulations, totaling *n* = 400 simulated subjects in each sample dataset. A total of 100 simulated datasets were generated for replicability under an overfitted model of *K* = 50 clusters under the 3 previously described approaches in 3.1. All analysis was performed using MATLAB 2021a.

### 3.3. Simulation Results

Model diagnostics indicated good mixing and successful convergence of model parameters across all three methods. Derived patterns were identified by setting the modal response to be the categorical level of each exposure variable that had the highest posterior probability of consumption. As illustrated in Figure 1, each of the methods successfully identified the true number of patterns (*K* = 3) as well as the modal response patterns in case A. The additional noise incorporated in case B generated additional clusters containing redundancies to the true patterns when using the unweighted method, but these were small in size (*π*_4_ *<* 0.03). Bias and precision in the expected prevalence of the patterns did differ across the three methods, as illustrated in Figure 2. The prevalence estimates for the unweighted method are clearly biased, whereas the estimates of the wtOLCM method show the least amount of bias, with slight sensitivity to noise for case B. Under the unweighted method, the MSE of the true pattern prevalence in the population was 0.015 and 0.016, respectively. Both the WFPBB and the wtOLCM methods had an improved estimation of the population prevalence compared to the unweighted case. Among all methods, wtOLCM had the best coverage of the true population prevalence 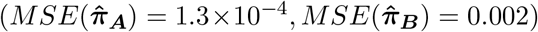 in both simulation cases.

**Fig. 1.**
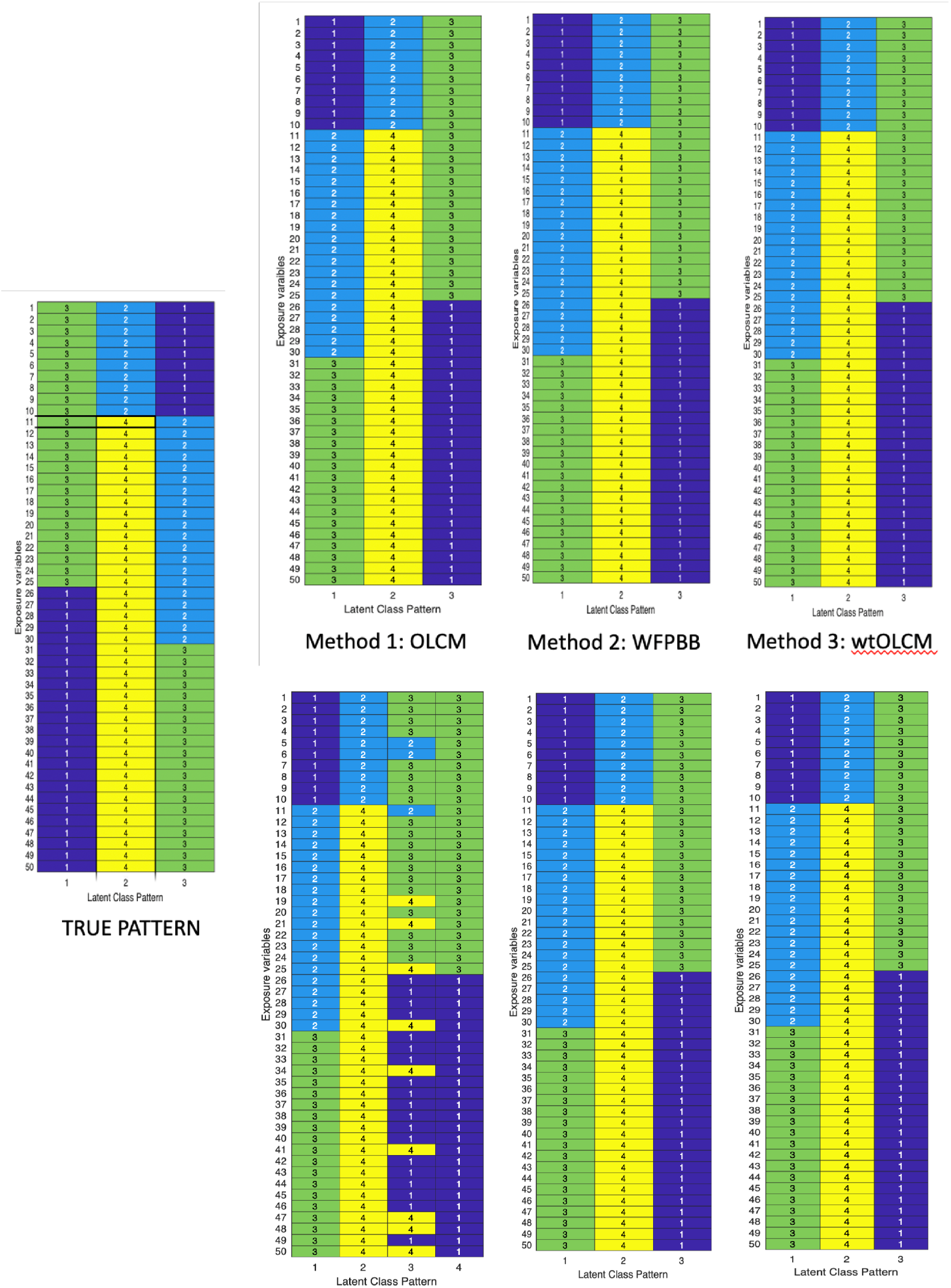
Modal consumption patterns identified from respective models compared to truth. Method 1: overfitted latent class model, ignoring weights; Method 2: weighted finite population Bayesian bootstrap; Method 3: weighted overfitted latent class model. Top indicates pattern under simulation case A. Bottom indicates pattern under simulation B. The additional noisy cluster is illustrated in method 1, where the size of this pattern had a prevalence of 0.02

**Fig. 2.**
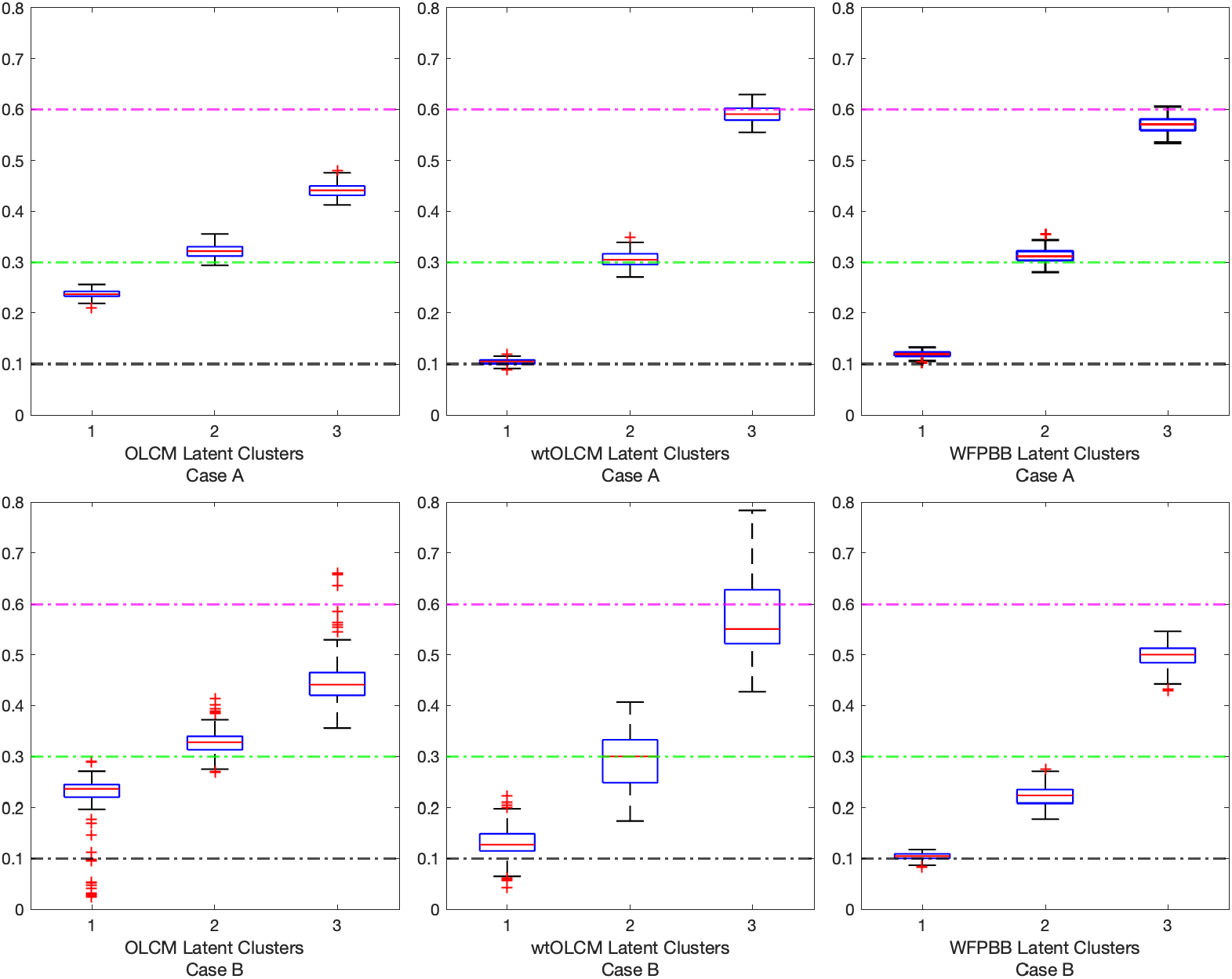
Predicted population prevalence from unweighted and weighted estimation approaches. Expected prevalence for each respective cluster is 0.1 (black), 0.3 (green), 0.6 (magenta)

## 4. National Health and Nutrition Examination Survey (NHANES)

The National Health and Nutrition Examination Survey (NHANES) is a population-based survey designed to assess the health and nutritional status of adults and children in the United States. The survey samples at least 9,000 people across various socioeconomic status (SES) levels each year residing in 15 randomly selected counties in the United States. Starting in 2011, NHANES created more granularity to the race/ethnicity variable, separating Mexican-American from Other Hispanic participants, as well as adding an identifier for Non-Hispanic Asian. For the scope of this study, we limited analysis to survey cycles containing the seven race/ethnicity groups, and adults aged 20 and over. Low-income participants were identified as those reporting at or below the 130% poverty income level.

Dietary intake was collected via the ‘What We Eat in America’ survey component of NHANES. Food items and beverages were consumed and recorded via two 24-hour recalls. Nutrients comprising these reported food/beverage items were calculated using the Food and Nutrition Database for Dietary Studies (FNDDS) and then converted into food pattern equivalents per 100 g of consumption based on the Dietary Guidelines for Americans (Committee et al., 2015; Bowman et al., 2016, 2017, 2018).

Dietary consumption data were summarized as 29 food groups and pooled across four NHANES survey cycles: 2011-2012, 2013-2014, 2015-2016, and 2017-2018. Consumption levels were derived by segmenting the data into no consumption (none=0%) and tertiles of positive consumption (Liu et al., 2019; Sotres-Alvarez et al., 2013). NHANES dietary weights were adjusted for the pooled survey years in accordance with protocols outlined in NHANES analytic guidelines (National Center for Health Statistics and Surveys, 2018; Chen et al., 2020).

Demographic information of the low-income adult participants collected in NHANES are detailed in Supplementary Table 2. The low-income sampled population reflected a demographic with the larger proportion of participants identifying as non-Hispanic White (47.6%), female (54.5%), and between 20-34 years old (35.7%). This sampled population reported an Alternative Healthy Eating Index (AHEI-2015) score of 49.2 out of 100, which is less than the overall national average of 58 out of 100. The mean Framingham 10-year risk score indicated a low risk of a CVD outcome occurring in the next ten years (*FRS* = 7.7).

### 4.1. NHANES Application of Weighed Overfitted Latent Class Model

#### 4.1.1. Fitting the Model

For our model, the normalization constant (*κ* = 9.79 *×* 10^3^) was calculated based on the sum of the sampled weights divide by the total sample size (*n* = 7561). We overfit the model with *K* = 50 latent classes. Estimation was performed using a Gibbs sampler of 10,000 iterations after a 15,000 burn-in and a thinning every 5 iterations. Posterior median estimates were derived from the MCMC output results. Flat, symmetric Dirichlet priors were fit with the probability of class assignment, ***π***, and the food item probability of consumption given assignment to specific latent class, ***θ***_***j·***|***k***_, *j* ∈ {1, …, *p*}, *k* ∈ {1, …, *K*}. A random permutation sampler was implemented to encourage mixing (Frühwirth-Schnatter, 2001). Dietary weights were calibrated and normalized for inclusion in analysis. We defined hyperprior 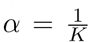 to conservatively moderate the rate of cluster growth as suggested in Rousseau and Mengersen (2011).

A common consequence in mixture modeling under Bayesian estimation is label switching, where label components swap assignment of individuals while the likelihood remains invariant (Stephens, 2000). We resolved this phenomenon by performing hierarchical clustering on a similarity matrix of size *n* × *n*. Matrix elements contained pairwise posterior probabilities of two subjects being clustered together in each MCMC iteration (Krebs, 1989; Medvedovic and Sivaganesan, 2002). Labels were identified based on subjects that remained clustered together through the sampling algorithm. Nonempty clusters were defined as any cluster containing at least 5% of the sampled participants. Dietary patterns were defined by identifying the consumption level corresponding to the highest posterior median probability for each food item in the set.

All data included for this study and code to reproduce the derived dataset and perform subsequent analyses are made available on the author’s GitHub repository: http://www.github.com/bjks10/NHANES_wtofm. Dietary data was originally obtained from the NHANES website (https://www.n.cdc.gov/nchs/nhanes) and processed in SAS 9.4. Statistical analysis and figures were performed in MATLAB 2021a. Posthoc analysis and table summaries were generated in R version 4.0.2.

#### 4.1.2. wtOLCM Results

The weighted overfitted latent class model identified five nonempty clusters in the low-income adult population. Figure 3 illustrates the posterior mean estimates of the probability of no consumption or high consumption given membership to a given dietary pattern. From this figure, we can see which foods were strongly favored to be consumed for various patterns. The very low probabilities of no consumption across all patterns for refined grains, oils, solid fats, and added sugar imply a general nonzero consumption by all low-income adults.

**Fig. 3.**
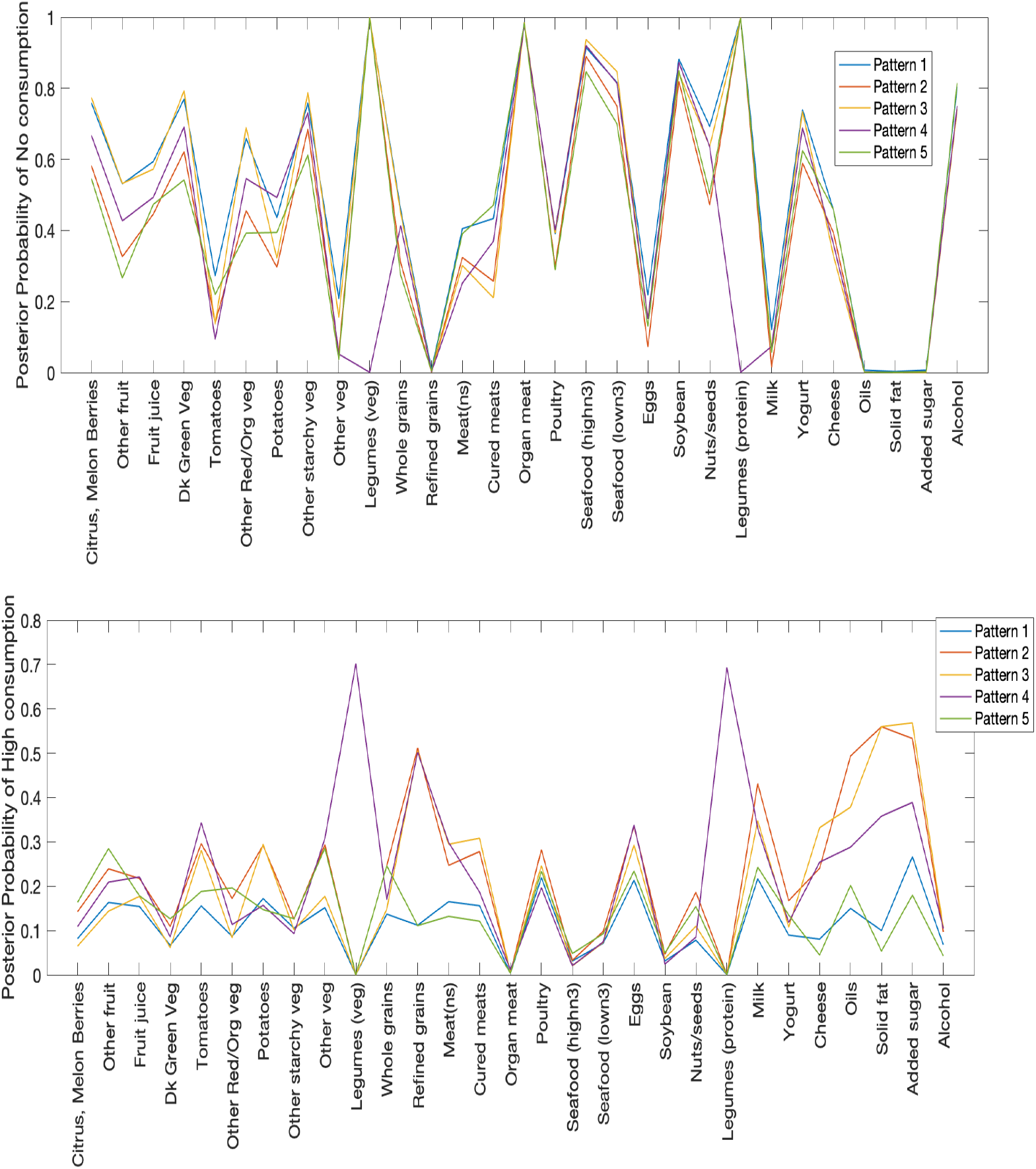
Low-income Adult population: (top) Posterior mean probability of no consumption of a given food item given membership in a specified pattern; (bottom) Posterior mean probability of high consumption of a given food item given membership in a specified pattern

Foods such as poultry, seafood, eggs, soybean and alcohol shared similar consumption behaviors across all diet patterns, but other foods differed by pattern. For example, patterns 1 and 5 had the lowest probabilities for consumption of cheese, oils, added sugars, and fats at the high consumption level. Patterns 2 and 3 had the highest probabilities for consumption of refined grains, potatoes, cured meats, oils, solid fat, and added sugar at the high consumption level. Lastly, pattern 4 distinctly had the highest probability of legumes being consumed at the high consumption level. Dietary pattern 1, followed by pattern 3, had the highest probabilities of no consumption of most fruits and vegetables.

Comparing more closely the posterior modes of consumption for each dietary pattern, we note that 15 foods shared a mode of non-consumption (i.e., consumption value of 1) across the five dietary patterns (Figure 4): citrus/melon/berries, fruit juice, dark green vegetables, other red/orange vegetables, potatoes, other starchy vegetables, whole grains, organ meat, poultry, seafood (high-n3), seafood (low-n3), soybean, nuts/seeds, yogurt, and alcohol. Pattern 1 showed strong similarities with Pattern 5. However, pattern 5 had comparatively higher levels of consumption of other fruit and milk. Patterns 2 and 3 also shared similar consumption of foods, with differences noted in the higher level of consumption for eggs and cheese in pattern 3. As previously noted, pattern 4 was the most distinguishable amongst the five patterns, with a high level of consumption favored in tomatoes, legumes (veg and protein), and non-specified meat.

**Fig. 4.**
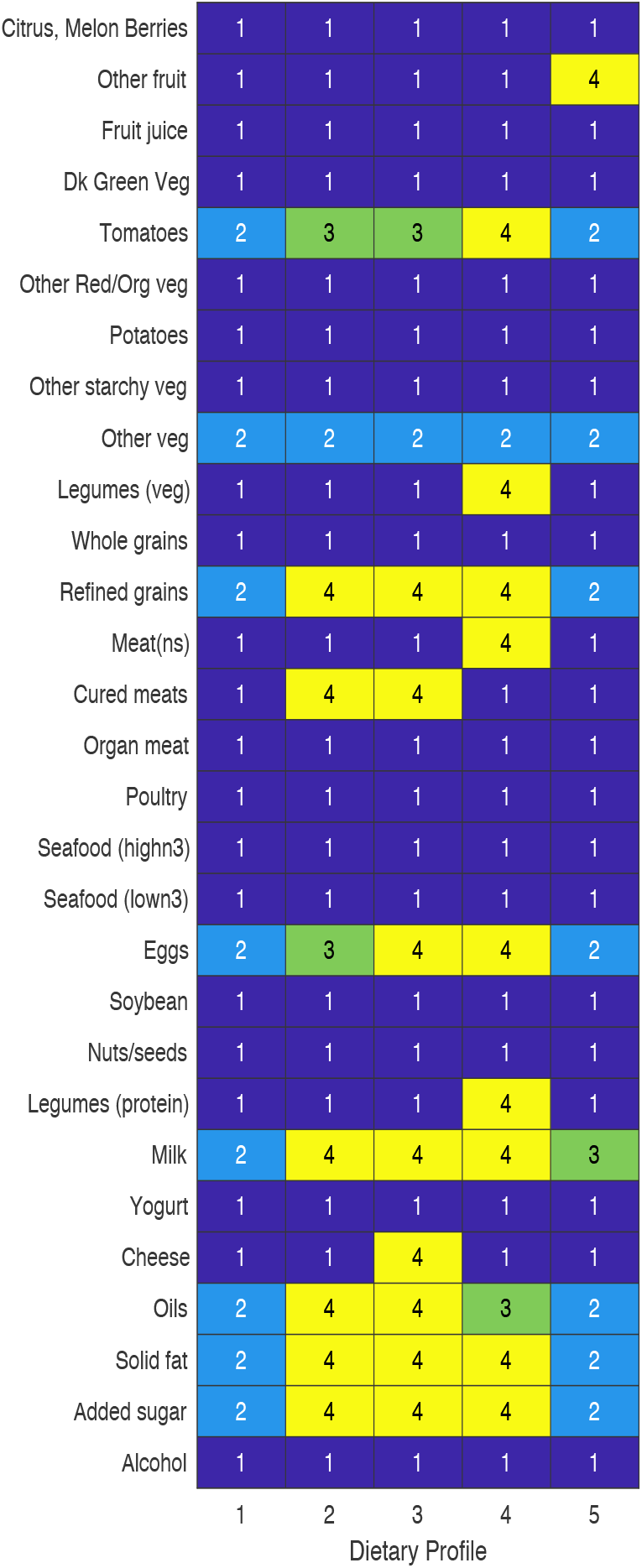
Posterior mode of consumption pattern of dietary patterns for non-incarcerated adults living at or below the 130% poverty level. Numbers represent levels of consumption: 1= None, 2=Low, 3=Medium, 4=High

Table 1 provides a summary of the demographics for participants assigned to each dietary pattern. Amongst the low-income adult population, participants assigned to pattern 5 had the highest average HEI-2015 score (57.4*±*0.6). This pattern favored a high consumption of other fruit, but a low consumption of refined grains and no consumption of meats. Pattern 3 had the lowest average HEI-2015 score (41.1±0.3). This pattern favored a high consumption of refined grains, cured meats, eggs, cheese, fats, oils, and sugars. Demographically, we observe that those in pattern 5 were predominantly male adults, whereas those assigned to pattern 3 were predominantly female adults. While non-Hispanic White participants held the majority of all patterns in our model, pattern 4, which uniquely favored a high consumption of legumes, was the only pattern where minority participants had a higher representation.

#### 4.1.3. Comparison to unweighted model

Ignoring the weights in the survey data generates different pattern results and prevalence. The cohort-specific model generated six clusters, ranging in size of 8.5% (unweighted pattern 3) to 30% (unweighted pattern 5). Consistent with what we saw in the simulation case, the OLCM of the cohort sample had similar patterns with the addition of a new pattern that looks like a mix of two separated patterns in the weighted model. A comparison of the consumption modes to describe each diet pattern for each model is provided in Supplementary Figure 2. About 97% of the cohort participants that were assigned to the largest pattern in the unweighted model also contributed to the largest pattern of the weighted model. Yes the consumption modes describing the respective patterns differed for four foods: non-specified meats, oils, solid fat, milk. For example, in the unweighted model, there was a 44% probability of not consuming non-specified meat for unweighted pattern 5 compared to 25% for weighted pattern 4. The smallest pattern prevalence identified in the unweighted model (pattern 3) differed in consumption of two foods from the weighted model (pattern 2): cured meats and tomatoes. These difference further highlight the consequence of applying survey data without the weights to appropriately estimate the true population, as opposed to the study cohort.

**Table 1.** Demographic distribution of Low-income Dietary patterns

|  | Pattern 1 |  | Pattern 2 |  | Pattern 3 |  | Pattern 4 |  | Pattern 5 |  |
| --- | --- | --- | --- | --- | --- | --- | --- | --- | --- | --- |
|  | Mean | SE | Mean | SE | Mean | SE | Mean | SE | Mean | SE |
| Overall | 21.7 | 0.6 | 11.7 | 0.6 | 22.0 | 0.7 | 29.3 | 1.0 | 15.4 | 0.9 |
| Race/Ethnicity |  |  |  |  |  |  |  |  |  |  |
| Mexican | 10.2 | 1.7 | 12.2 | 1.6 | 11.0 | 1.5 | 26.4 | 2.9 | 10.8 | 1.5 |
| Other Hispanic | 8.4 | 1.2 | 7.5 | 1.2 | 5.2 | 0.9 | 15.7 | 1.8 | 7.9 | 1.0 |
| Non-Hispanic White | 50.5 | 2.9 | 54.3 | 3.4 | 55.1 | 2.7 | 37.0 | 2.8 | 47.9 | 3.2 |
| Non-Hispanic Black | 21.2 | 2.1 | 18.2 | 2.1 | 22.1 | 2.6 | 11.1 | 1.2 | 16.8 | 1.8 |
| Non-Hispanic Asian | 5.4 | 0.9 | 4.5 | 0.8 | 1.2 | 0.2 | 5.8 | 0.9 | 12.5 | 1.8 |
| Mixed/Other | 4.4 | 1.0 | 3.3 | 0.7 | 5.5 | 0.7 | 4.0 | 0.8 | 4.2 | 0.9 |
| Gender |  |  |  |  |  |  |  |  |  |  |
| Male | 63.2 | 1.5 | 47.2 | 2.5 | 38.6 | 1.5 | 53.6 | 1.5 | 72.2 | 2.1 |
| Female | 36.8 | 1.5 | 52.8 | 2.5 | 61.4 | 1.5 | 46.4 | 1.5 | 27.7 | 2.1 |
| Age Group |  |  |  |  |  |  |  |  |  |  |
| 20-34 years | 33.1 | 1.9 | 38.2 | 2.8 | 46.6 | 2.5 | 35.2 | 2.1 | 22.8 | 2.0 |
| 35-49 years | 24.9 | 1.5 | 20.5 | 1.6 | 25.6 | 1.5 | 26.6 | 1.4 | 20.0 | 1.5 |
| 50-64 years | 26.1 | 1.4 | 22.5 | 2.0 | 18.2 | 1.6 | 25.3 | 1.7 | 27.3 | 2.1 |
| 65+ years | 15.9 | 1.1 | 18.8 | 2.1 | 9.6 | 1.0 | 12.9 | 1.0 | 30.0 | 2.1 |
| HEI 2015 Score | 45.8 | 0.5 | 50.3 | 0.6 | 41.1 | 0.3 | 53.2 | 0.5 | 57.4 | 0.6 |
| Framingham 10YR Risk | 7.7 | 0.3 | 7.6 | 0.4 | 7.1 | 0.4 | 7.4 | 0.3 | 9.3 | 0.5 |
| CVD Risk factors |  |  |  |  |  |  |  |  |  |  |
| Hypertension | 34.9 | 2.1 | 30.9 | 3.5 | 30.9 | 2.0 | 30.2 | 1.7 | 36.4 | 2.8 |
| Hypercholesteremia | 76.7 | 1.9 | 67.9 | 4.2 | 69.4 | 1.9 | 75.5 | 1.9 | 68.1 | 2.5 |
| Obesity | 45.7 | 2.6 | 42.2 | 3.8 | 41.4 | 2.5 | 40.1 | 1.9 | 38.0 | 2.7 |
| Diabetes | 12.8 | 1.0 | 10.0 | 1.6 | 8.2 | 1.2 | 9.1 | 1.1 | 14.2 | 1.6 |
| Smoker | 34.5 | 3.1 | 18.6 | 2.5 | 34.9 | 3.0 | 19.2 | 2.1 | 11.9 | 1.8 |

## 5. Discussion

The weighted overfitted latent class model (wtOLCM) for survey data, proposed in this paper, is an extension of the standard latent class model and integrates a Bayesian non-parametric survey-weighted approach to account for sampling variability in its parameter estimation. Our simulation study compared our proposed model with other standard approaches used for surey data. The results showed that wtOLCM had an improved estimation of the true population prevalence as well as pattern identification, particularly as more heterogeneity is introduced. We applied our model to dietary survey data collected in the 2011-2018 National Health and Nutrition Examination Surveys in order to better examine the dietary patterns of US adults living at or below the 130% poverty income level. Our model identified five dietary patterns in this sampled subset. Application of our model to this target population allowed us to leverage survey weights to obtain representative estimates from a smaller, often underrepresented and understudied, subset of the surveyed participants. Ignoring the weights provided by the survey would have biased our results and led to misleading inference of this low-income adult population. This method builds its strength on its generalizability and use in nationally representative dietary surveys, yet recognizes the concerns of overgeneralization. Dominating demographics can still influence pattern distribution in a given population. Non-Hispanic White participants have historically dominated surveys and studies that examine diet-disease relationships (Ohlhorst et al., 2013; Fahlman et al., 2010). This underrepresentation of minority subgroups makes it difficult to identify a uniquely separate cluster under the global clustering assumption, if the derived pattern is not shared amongst all individuals. An overrepresentation of this subgroup can mask accurate pattern identification for racial/ethnic minorities who may be at greatest risk of chronic disease. This is exemplified in our model through Pattern 4, which had the most distinguishable dietary consumption pattern. Compared to the other five patterns, this contained the smallest proportion of non-Hispanic White adults, but still the largest relative proportion amongst the other racial/ethnic subgroups. If certain subgroups are important to understand nutrition disparities, those subgroups should be studied in a separate analysis or a more advanced method that is able to jointly account for subgroup differences should be implemented. To our knowledge a few advanced methods have been used to better examine subpopulation behavior differences, but incorporating the complex survey design directly into model estimation has not yet been fully explored (De Vito et al., 2019; Stephenson and Willett, 2022).

While the utility of this model has effectively demonstrated its use in diet survey data, we must also acknowledge that the dietary intake data used is limited by its reliance on self-reporting. Several nutrition studies have found that prudent foods like vegetables and fruits are often overreported and less prudent foods like fats and oils are frequently underreported (Haraldsdó, 1993; Amanatidis et al., 2001). These tendencies to misreport have been associated with demographics such as BMI, age, sex, socioeconomic status, as well as other psychosocial and cognitive factors (Poslusna et al., 2009; Klesges et al., 1995; Tooze et al., 2004).

Methods such as doubly labeled water and biomarkers for select nutrients are available to validate dietary assessment tools, but these instruments are beyond the scope of tools utilized in the National Health and Nutrition Examination Survey. In spite of this limitation, the misreporting rate remains relatively low and the instruments can still be deemed relatively reliable (Tooze et al., 2004; Yuan et al., 2017). Another limitation of dietary recalls is the inability to capture day-to-day variation. As a result, these dietary patterns are based on one or two days of dietary records, which may or may not reflect participants’ regular dietary behaviors. Alternative dietary assessments, such as food frequency questionnaires and 7-day daily diet records, are available to capture more episodic and rarely consumed foods. However, more detailed assessments are often costly and seldom widely available in large population-based surveys. Future research can explore ways to integrate these tools, when available, to quantify the unknown variation and uncertainty that comes from misreporting in dietary assessments.

The clustering approach applied in this paper, as well as more traditionally used cluster and factor analysis, are all generated independent of any health outcome. Yet, when looking at exposures from a multi-dimensional perspective, these exposures may be driven by an underlying health outcome, in which case a more supervised approach may yield more useful information to understand how the combination of these exposures (e.g., dietary habits) can drive a known outcome (e.g., cardiometabolic health). In addition, this paper did not report estimates of variance and uncertainty for the wtOLCM. Variance estimates are expected to exhibit slightly less than nominal coverage. Though methods have been proposed to address this issue (Williams and Savitsky, 2021; León-Novelo and Savitsky, 2019). Incorporation of these methods into a model-based clustering framework, such as the wtOLCM, remains an area of active research. Further research is needed to develop supervised clustering methods that address the issue of confounding overgeneralizations and are applicable in population surveys with complex survey designs.

## 6. Software Availability Statement

Software in the form of MATLAB code, together with a sample input data set and complete documentation, is available in the GitHub repository https://github.com/bjks10/NHANES_wtofm.

## Data Availability

All data produced are available online at GitHub repository: https://github.com/bjks10/NHANES_wtofm

## Acknowledgements

The authors are grateful to Walter Willett, DC Rao, and Lei Liu for helpful comments on earlier versions of this work. This study was supported in part by NHLBI grant R25 HL105400 to DC Rao and Victor G. Davila-Roman.

