## Supplementary Materials for "Identifying Dietary Consumption Patterns from Survey Data: A Bayesian Nonparametric Latent Class Model"

### SUPPORTING WEB MATERIALS FOR IDENTIFYING DIETARY CONSUMPTION PATTERNS FROM SURVEY DATA: A BAYESIAN NONPARAMETRIC LATENT CLASS MODEL

STEPHENSON ET AL.

Contents

1. Weighted Finite Population Bayesian Bootstrap method 1
References 2
2. Posterior computation of weighted overfitted latent class model 2
3. Supplementary Tables 3
4. Supplementary Figures 3

#### 1. Weighted Finite Population Bayesian Bootstrap method

In the weighted finite population Bayesian bootstrap (WFPBB) method, the population from which the survey data arise is assumed to be finite. A pseudo-representative sample (PRS) approximates a simple random sample from the finite population. To generate a PRS, an "unweighted" synthetic population is first created using a weighted multiple imputation approach, where non-sampled units in the finite population are viewed as missing data and are imputed by sampling from the posterior predictive distribution given the sampled data and the survey weights. Imputation proceeds via the WFPBB method (Dong et al., 2014; Cohen, 1997), which extends the Bayesian bootstrap (Rubin, 1981) and its weighted version (Lo, 1993) to also account for a finite population. Dong et al. (2014) show that sampling from the posterior predictive distribution of the non-sampled units given the sampled data and survey weights is equivalent to sampling from a weighted Pólya urn scheme. This allows for simple implementation of the procedure to generate the synthetic population. A total of 200 synthetic populations are generated, and the corresponding 200 PRS's are formed by taking a simple random sample without replacement from a synthetic population.

Posterior inference is obtained by running the unweighted overfitted latent class model on each PRS as if it were the actual sampled data, and then combining results across the samples. Parameter point (and variance) estimates are obtained by taking the mean of the posterior median (and variance) estimates across all PRS's. One notable limitation of this procedure is that it does not allow for classification of individuals into clusters. Incorporation of survey weights into Bayesian clustering methods that allow for classification within a multiple imputation framework is a potentially promising direction for future research.


#### 2. Posterior computation of weighted overfitted latent class model

(1) Update $z_i$ for $i \in (1, \dots, n)$:

$$Pr(z_i = k) = \frac{\pi_h \prod_{j=1}^{p} \prod_{c=1}^{d_j} \theta_{jc|h}^{1(y_{ij}=c)}}{\sum_{k=1}^{K} \pi_k \prod_{j=1}^{p} \prod_{c=1}^{d_j} \theta_{jc|k}^{1(y_{ij}=c)}}$$

(2) Update $\pi = (\pi_1, \dots, \pi_K)$:

$$\pi_{1:K} \sim \text{Dir}\left( \alpha + \sum_{i=1}^{n} \mathbf{1}(z_i = 1) \times \frac{w_i}{\kappa}, \dots, \alpha + \sum_{i=1}^{n} \mathbf{1}(z_i = K) \times \frac{w_i}{\kappa} \right),$$

(3) Update $\theta_{j\cdot|h} = (\theta_{j1|k}, \dots \theta_{jd|k})$ for all $j \in (1, \dots, p), k \in (1, \dots, K)$:

$$\theta_{j\cdot|k} \sim \text{Dir}_{d_j} \left( \eta + \sum_{i:z_i=k} \mathbf{1}(y_{ij} = c | z_i = k) \times \frac{w_i}{\kappa} \right)$$

TABLE 1. Distribution of patterns across simulated population containing four subpopulations and three uniquely distinct patterns

| | **Pattern 1** | **Pattern 2** | **Pattern 3** | **Total** |
|---|---|---|---|---|
| Subpopulation 1 | 25 | 900 | 75 | 1000 |
| Subpopulation 2 | 0 | 175 | 1825 | 2000 |
| Subpopulation 3 | 25 | 425 | 1050 | 1500 |
| Subpopulation 4 | 450 | 0 | 50 | 500 |
| Total | 500 | 1500 | 3000 | 5000 |

TABLE 2. NHANES 2011-2018 adult participant demographics

| | Overall | | | Low-income | | |
|---|---|---|---|---|---|---|
| Demographics | N | % | (SE) | N | % | (SE) |
| **Race/Ethnicity** | | | | | | |
| Mexican | 2632 | 9.2 | (1.0) | 1353 | 15.4 | (1.7) |
| Other Hispanic | 2026 | 6.3 | (0.6) | 950 | 9.7 | (1.0) |
| Non-Hispanic White | 7480 | 64.0 | (1.7) | 2390 | 47.6 | (2.3) |
| Non-Hispanic Black | 4471 | 11.3 | (1.0) | 1963 | 17.4 | (1.5) |
| Non-Hispanic Asian | 2282 | 5.8 | (0.5) | 609 | 5.6 | (0.7) |
| Mixed/Other | 716 | 3.4 | (0.3) | 296 | 4.3 | (0.5) |
| **Gender** | | | | | | |
| Male | 10077 | 48.8 | (0.5) | 3517 | 45.5 | (0.8) |
| Female | 9530 | 51.2 | (0.5) | 4044 | 54.5 | (0.8) |
| **Age** | | | | | | |
| 20-34 Years | 4991 | 28.9 | (0.8) | 2129 | 35.7 | (1.5) |
| 35-49 Years | 4801 | 25.5 | (0.7) | 1767 | 24.3 | (0.9) |
| 50-64 Years | 5246 | 26.9 | (0.6) | 1976 | 23.9 | (0.9) |
| 65+ Years | 4569 | 18.8 | (0.6) | 1689 | 16.1 | (0.8) |
| AHEI2015 Score | 14865 | 51.6 | (0.3) | 5823 | 49.2 | (0.3) |
| Framingham 10YR Score | 18226 | 8.1 | (0.1) | 6904 | 7.7 | (0.2) |
| **CVD Risk factors** | | | | | | |
| Hypertension | 8260 | 32.6 | 0.9 | 2446 | 32.3 | 1.0 |
| Obesity | 8198 | 39.0 | 0.9 | 2589 | 41.4 | 1.2 |
| Diabetes | 2010 | 9.3 | 0.4 | 643 | 10.6 | 0.6 |
| High Cholesterol | 9906 | 75.1 | 0.8 | 2874 | 72.4 | 1.0 |
| Smoker | 3487 | 15.1 | 0.7 | 1605 | 24.4 | 1.4 |

#### 3. SUPPLEMENTARY TABLES

#### 4. SUPPLEMENTARY FIGURES

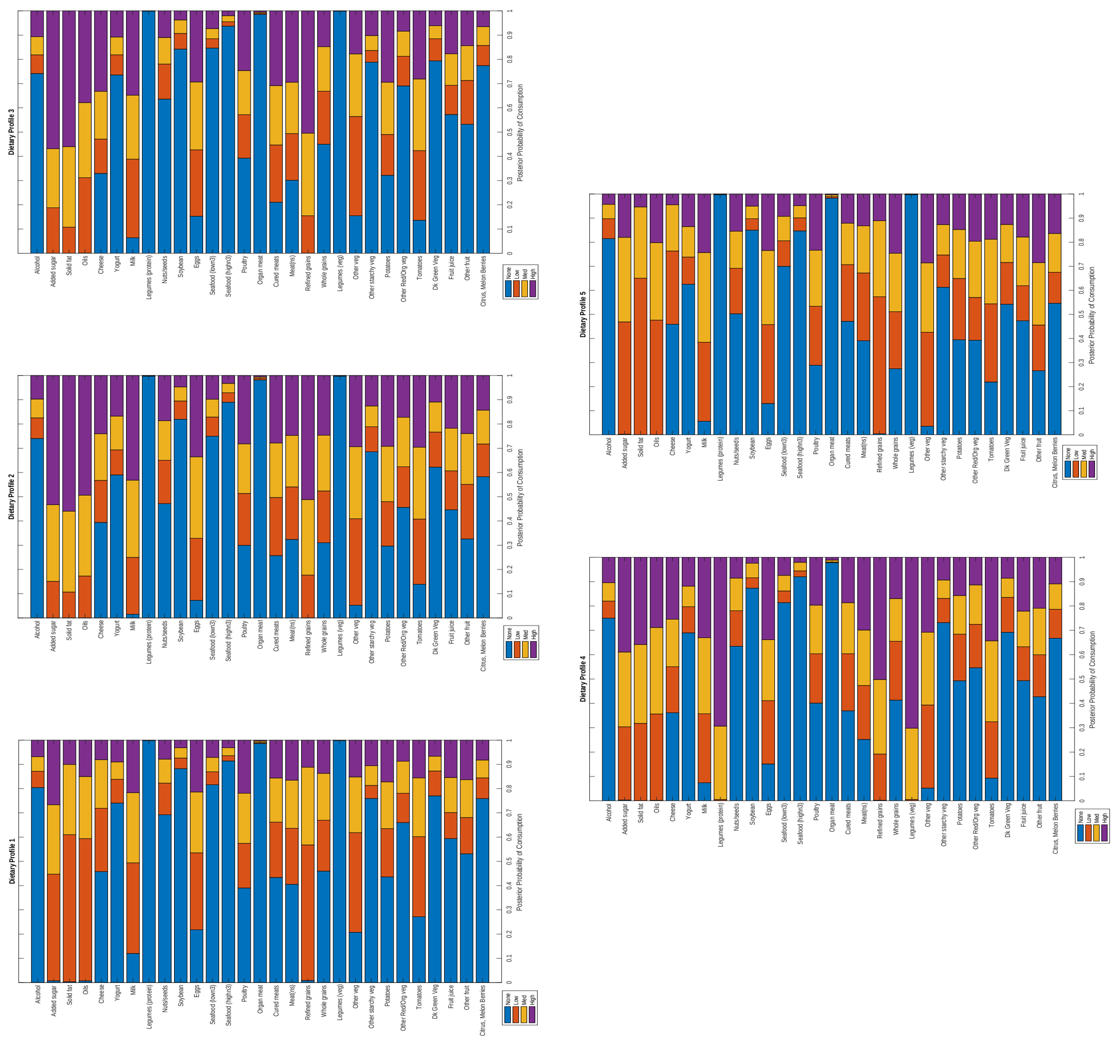


Figure 1. Distribution of dietary consumption for each dietary pattern derived from NHANES data on Low-income adult population

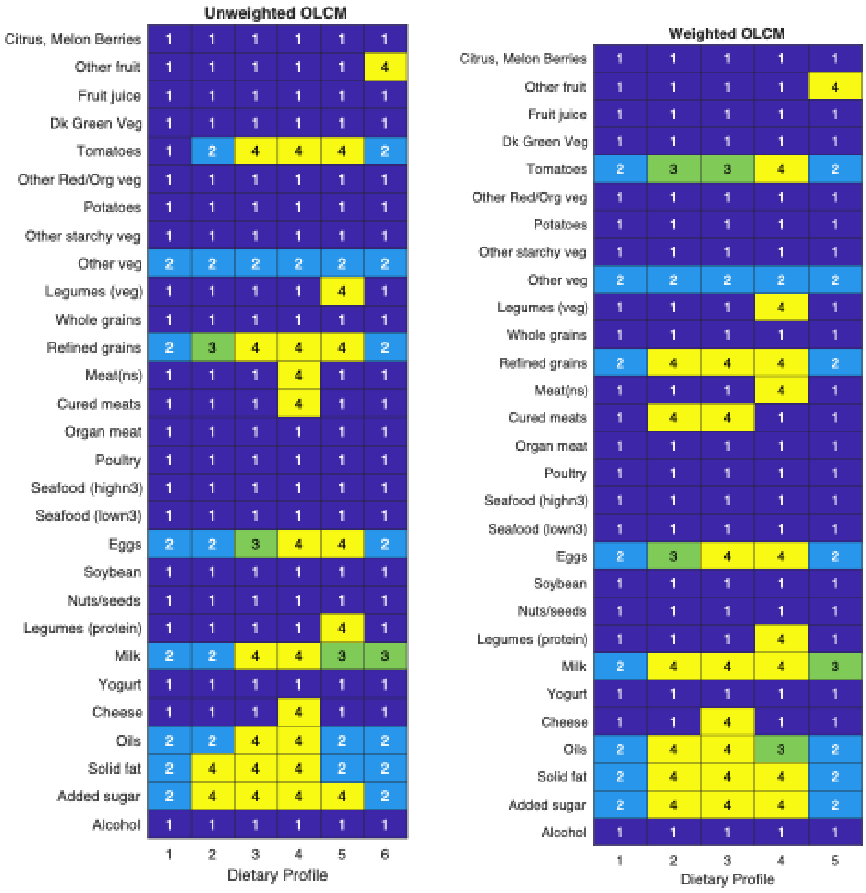


FIGURE 2. Posterior mode of consumption pattern of dietary patterns for non-incarcerated adults living at or below the 130% poverty level. Left=unweighted model with ($\pi_{OLCM}$ = $0.18, 0.13, 0.09, 0.16, 0.30, 0.16$). Right = weighted model with ($\pi_{wtOLCM}$ = $0.22, 0.12, 0.22, 0.29, 0.15$). Numbers represent levels of consumption: 1= None, 2=Low, 3=Medium, 4=High.
